# Procoagulant plasma in three individuals with stabilin-2 deficiency

**DOI:** 10.64898/2026.10.04.26363616

**Authors:** Rüdiger Adam, Alexander J. Jordan, Taras Valovka, Carla S. M. Seidl, Karin Kreidl, Carl-Erik Dempfle, Dorota Garczarczyk-Asim, Annelie Siegemund, Stefanie Nittka, Robert Müller, Petra Bürgisser, Ruben Bierings, Christoph Brochhausen-Delius, Krystin Krauel, Alice Assinger, Lea Berger, Eray Yagmur, Anja Schmaus, Werner Streif, Cyrill Géraud, Thomas Müller, Andreas R. Janecke

**Author notes:** Equal contribution (Cyrill Géraud, Thomas Müller, Andreas R. Janecke). Correspondence: Andreas R. Janecke, M.D. & Thomas Müller, M.D., Medical University of Innsbruck, Department of Pediatrics I, Anichstrasse 35, A-6020 Innsbruck, Austria; &, & Cyrill Geraud, European Center for Angioscience and Section of Clinical and Molecular Dermatology, Department of Dermatology, Venereology, and Allergology, University Medical Center and Medical Faculty Mannheim, Heidelberg University, 68167 Mannheim, Germany.

## Abstract

**BACKGROUND:** Rare *STAB2* loss-of-function variants emerged in population studies as being associated with elevated venous thromboembolism (VTE) risk. *STAB2* encodes the cell-surface scavenger receptor stabilin-2, and reduced stabilin-2 function might increase VTE risk via elevated plasma levels of its ligands, including von Willebrand factor (VWF) and coagulation factor VIII (FVIII).

**METHODS:** An adolescent patient with dependency on parenteral nutrition had experienced episodes of central venous catheter-related thrombosis, including the complete thrombotic occlusion of the deep pelvic veins, and pulmonary embolism. Exome sequencing for increased VTE risk identified a homozygous *STAB2* splice-site variant and its effect on splicing was characterized in fibroblast-derived RNA and in a minigene splicing system. Plasma VWF, FVIII, and fibrinogen and serum hyaluronic acid (HA) levels were determined. Immunohistochemistry was performed on liver biopsy tissue.

**RESULTS:** The *STAB2* splice-site variant caused aberrant *STAB2* mRNA splicing. The patient’s healthy mother and one healthy sibling were also homozygous for the *STAB2* splice-site variant but had no history of VTE. Plasma FVIII levels and VWF activity were elevated in all three, and VWF antigen was elevated in two homozygous individuals. Strikingly, approximately 1000-fold elevated HA levels were observed in all three homozygous individuals. Stabilin-2 immunohistochemistry on a liver biopsy from the proband’s mother revealed no detectable signal, whereas control human liver showed strong labelling of liver sinusoidal endothelial cells. The *STAB2* splice-site variant thus results in loss of detectable stabilin-2 protein.

**CONCLUSIONS:** To current knowledge, this is the first family-based report of stabilin-2 deficiency with markedly elevated circulating HA, VWF, and FVIII levels in humans. These findings support impaired stabilin-2-dependent ligand clearance as a thrombosis susceptibility state with incomplete penetrance. The data are consistent with a significant role for stabilin-2 in circulating HA clearance in humans, but do not establish elevated HA itself as a cause of VTE.

## Introduction

Various hereditary and acquired risk factors for venous thromboembolism (VTE), encompassing deep vein thrombosis (DVT) and pulmonary embolism, have been identified, including elevated levels of von Willebrand factor (VWF) and coagulation factor VIII (FVIII).^1^ Variants in *STAB2* influence plasma levels of VWF and FVIII in individuals with and without VWF deficiency.^1–3^ Recently, a genome-wide screen of 393 patients and 6114 control subjects of European ancestry using exome sequencing reported rare pathogenic variants in *STAB2* that were associated with unprovoked VTE. Such putative *STAB2* loss-of-function (LoF) changes conferred an elevated risk of VTE with an odds ratio of 3.37.^4^ Subsequently, an elevated VTE risk for *STAB2* variants was also demonstrated in a Swedish population-based cohort.^5^

*STAB2* encodes the cell-surface scavenger receptor stabilin-2, which is expressed by sinusoidal endothelial cells in the liver, spleen, bone marrow, and lymphatics and comprises multiple ligand-binding domains.^6^ Human stabilin-2 can bind, internalize, and clear VWF and the VWF-FVIII complex,^7^ suggesting that *STAB2* LoF variants impair endothelial clearance of VWF and FVIII and thereby influence plasma levels and molecular-weight distribution of VWF as well as plasma levels of FVIII. Stabilin-2 has also been identified as a clearance receptor for serum hyaluronic acid (HA) in humans^8^ and mice,^9,10^ for other glycosaminoglycans,^11,12^ and for phosphatidylserine-positive erythrocytes.^13^ Moreover, an exome-wide association study and gene-level collapsing analysis of rare coding variants identified 41 protein associations in plasma protein clearance for *STAB2* in the UK Biobank.^14^

While *STAB2* variants have recently emerged as VTE risk alleles, discrimination between neutral and VTE-causing *STAB2* variants remains challenging, as does determining the extent of individual VTE risk elevation.^5^ Here, the phenotypic expression of a homozygous *STAB2* splice-site variant that results in absence of stabilin-2 protein is described in three members of one family.

## Materials and methods

The data that support the findings of this study are available from the corresponding author on reasonable request.

## Human subjects

Patients with suspected stabilin-2 deficiency and their family members were recruited through a proband who presented postnatally with one form of intractable diarrhea, microvillus inclusion disease (MVID). Primary and secondary hemostasis were assessed in detail; blood was drawn by phlebotomy into heparinized tubes for hemostaseology and into EDTA tubes for DNA extraction. All study participants provided written informed consent in accordance with the Declaration of Helsinki. The study was approved by the Ethics Committee of the Medical University of Innsbruck (number UN-4501), Innsbruck, Austria. Participants were not compensated.

## Exome sequencing and sequence data analysis

Genomic DNA was extracted from peripheral blood samples and buccal swabs using standard procedures. The proband’s coding DNA fragments were enriched with the Roche/Nimblegen SeqCap EZ v2.0 kit and captured libraries were sequenced on a HiSeq platform (Illumina, San Diego, CA) with 100-bp paired-end reads. Sequencing reads were aligned to the human reference genome GRCh37 with BWA.^15^ Single-nucleotide variant and insertion/deletion calls were performed using PICARD and SAMTOOLS.^15,16^ Copy-number variants were called with panelcn.MOPS software.^17^

Variant calling, filtering for allele frequency below 0.005 in gnomAD, and in silico evaluation for pathogenicity were performed as described.^18^ Variants predicting protein truncation and obtaining strong PolyPhen-2^19^ and SIFT^20^ scores were preferentially considered pathogenic or likely pathogenic.

## *STAB2* splice-site variant segregation and in silico splicing analysis

Sanger sequencing of a 275-bp genomic PCR fragment obtained with primers STAB2_55f 5’-CCTCCATGAGGGCACAGAC and STAB2_55r 5’-ACTGGACAACGAGCAGGAGC enabled segregation analysis within the proband’s family of a *STAB2* variant at the intron 54-exon 55 splice site detected by exome sequencing. A total of nine family members were available for segregation analysis. *STAB2* variant designation is based on the NCBI reference sequence for *STAB2*.

SpliceAI^21^ and Pangolin^22^ web tools were used to assess the effect of the *STAB2* splice-site variant on mRNA splicing.

## Fibroblast culture and *STAB2* transcript analysis

Primary human patient fibroblasts established from skin biopsies were available to analyze the transcript consequence of the canonical *STAB2* splice-site mutation. Fibroblasts were regularly tested for mycoplasma and maintained in Dulbecco’s Modified Eagle Medium supplemented with 10% fetal bovine serum, 100 U/mL penicillin, and 100 μg/mL streptomycin at 37 °C in a humidified atmosphere with 5% CO2.^23^ Total RNA was extracted using the Qiagen RNeasy Mini Kit from cells treated or untreated with puromycin at a final concentration of 200 μg/mL for 6 hours before harvest. cDNA was synthesized with the LunaScript RT SuperMix Kit (New England Biolabs).

Part of the coding sequence of *STAB2* was amplified using primers *STAB2*_cDNA_54f 5’-GAGTGTGGGAGCTGTGTCAA and *STAB2*_cDNA_56r 5’-ACAATCAGGCCCGAATCTC, followed by nested PCR using primers *STAB2*_cDNA_nest_54f 5’-CTGCCCAAGGTGGAGTAAAC and *STAB2*_cDNA_nest_56r 5’-ATTGAAGCCGGTGTTGCAT. The nested PCR product was sequenced using BigDye terminator chemistry on an ABI PRISM 3100 Genetic Analyzer. Data were analyzed with Sequencher software.

## Minigene construction and analysis

Because *STAB2* expression in fibroblasts was low and transcript detection required nested PCR, aberrant splicing was additionally assessed using a minigene approach. DNA fragments containing *STAB2* exon 55 and flanking intronic regions, either wild-type or mutant, were amplified from genomic DNA and cloned into the Exontrap vector pET01 (Mobitec, Goettingen, Germany) using standard restriction-ligation procedures. Wild-type and mutant minigenes were confirmed by sequencing and transfected into HEK293T cells. After 48 hours, total RNA was extracted using the Monarch Total RNA Miniprep Kit, reverse-transcribed with LunaScript RT SuperMix, amplified using vector-specific primers, and sequenced. Cell culture and transfection procedures were performed as described previously,^24^ and a structured, transparent, accessible, reproducible (STAR) protocol is available.^25^

## Stabilin-2 immunohistochemistry on liver biopsies

Immunohistochemistry was performed on formalin-fixed, paraffin-embedded tissue using a monoclonal anti-Stabilin-2 antibody (clone 3.1; InVivo Biotech Services, Berlin, Germany) on a BenchMark ULTRA automated stainer (Roche Diagnostics, Mannheim, Germany). Liver biopsy of the proband’s mother was performed for suspected hepatic steatosis in adulthood. Tissue from the proband’s mother and control liver tissue routinely included as a positive control in the diagnostic staining protocol was mounted on the same slide and processed in the same run. Pretreatment was carried out with Protease 1 for 8 min, and the primary antibody was applied at a dilution of 1:100. Detection was performed with the OptiView DAB IHC Detection Kit (Roche Diagnostics, Mannheim, Germany), followed by counterstaining with hematoxylin.

## Thrombin generation assay

Thrombin generation potential in a healthy individual homozygous for the *STAB2* mutation was obtained on ST Genesia (Diagnostica Stago, France), with and without thrombomodulin using the reagent ST Thromboscreen, and performed as described.^26,27^ Venous blood was collected into 3.2% trisodium citrate Monovette tubes.

Samples were centrifuged twice at 2500 g for 15 minutes at 20 °C immediately after collection, and plasma was stored at –80 °C until analysis.

## Measurement of HA serum concentrations

Blood samples for HA measurement were collected in the morning. After centrifugation at 2000 g, serum samples were stored at –80 °C until analysis. Quantitative determination of serum HA concentrations in the proband, family members, and controls was carried out using an automated latex agglutination assay according to the manufacturer’s instructions (Centaur XP, Siemens Healthineers, Forchheim, Germany). References and standard-curve calibration have been reported previously.^28,29^

## HA size distribution

The molecular size distribution of HA in patients with markedly elevated serum HA levels was assessed as described previously.^30^

## Blood viscosity analysis in humans and mice

To assess whether blood viscosity was altered by high levels of HA in the circulation, blood samples from stabilin-2-deficient individuals and from 10 stabilin-2-knockout and wild-type mice of both sexes were analyzed by cone-and-plate viscometry using a DVNext Wells-Brookfield Cone/Plate Rheometer.^31^ All animals received humane care in compliance with the Guide for the Care and Use of Laboratory Animals, and all experiments were approved by the animal ethics committee of Baden-Württemberg (Regierungspräsidium Karlsruhe).

## Results

The proband (II-1, Figure 1A) presented with intractable diarrhea at birth, and a diagnosis of MVID related to syntaxin-3 (*STX3*) deficiency was established through exome sequencing and functional testing. ADDIN EN.JS.CITE ^32^ Nutritional and fluid needs were sustained exclusively through continuous parenteral nutrition during early childhood. He later developed eight episodes of prolonged venous thrombosis, both catheter-related and independent of central venous access, as well as multiple central-line-related episodes of septicemia. Two thrombotic events resulted in complete occlusion of the deep pelvic veins. The proband was born to healthy, consanguineous South Asian parents; one younger brother is healthy, and another younger brother has MVID and requires parenteral nutrition.

**Figure 1.**
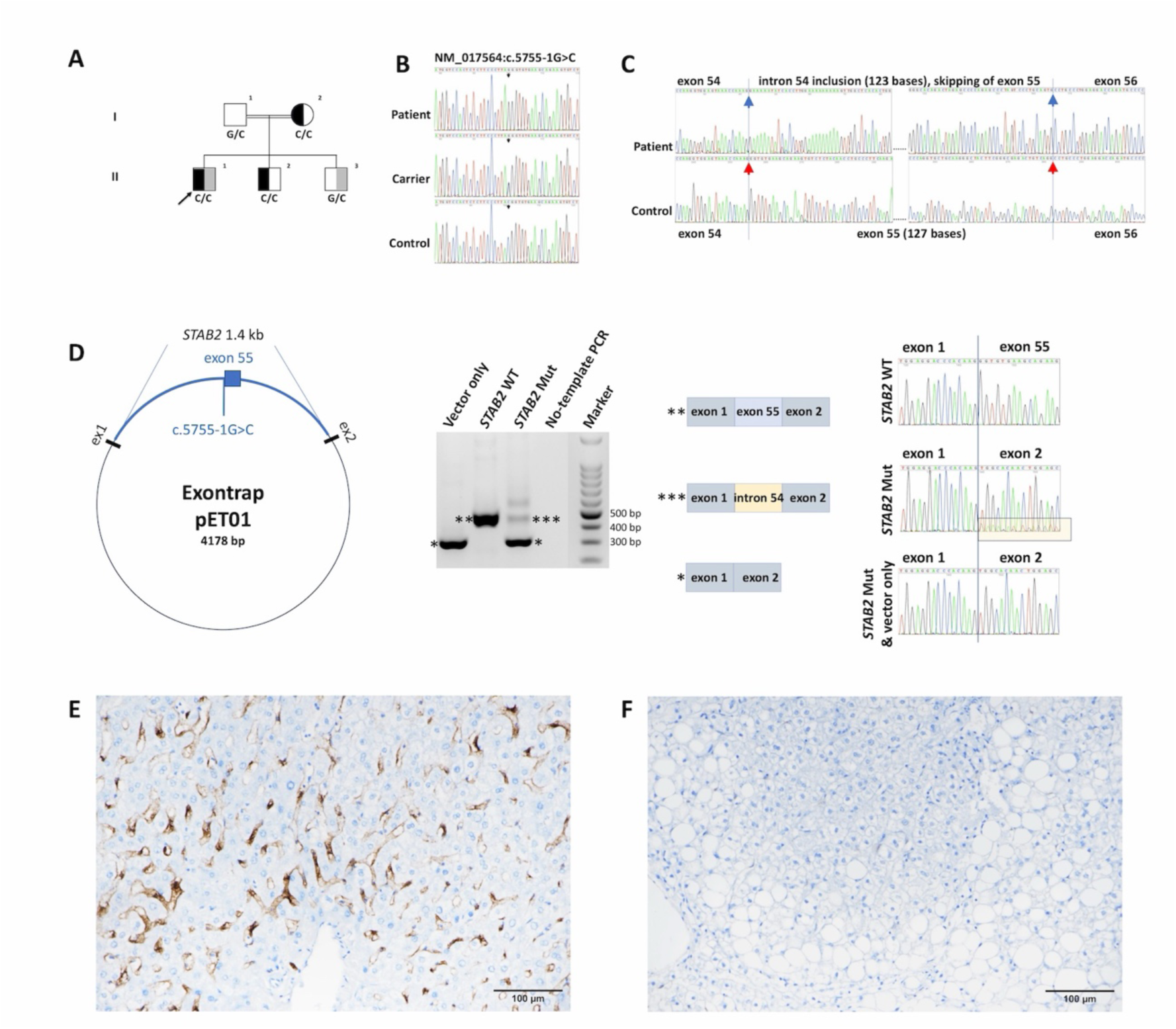
A *STAB2* splicing defect resulting in stabilin-2 deficiency in one family. **A.** Family tree and segregation of the *STAB2* splice-site variant NM_017564:c.5755-1G>C (rs570260501) with elevated VWF and FVIII plasma levels and markedly elevated serum HA levels, denoted by black shading of individuals’ symbols. Syntaxin-3-related MVID in the family is denoted by grey shading of symbols; the proband is marked by an arrow. To protect participant privacy, the pedigree is restricted to the proband’s parents and siblings; data on additional relatives are available from the corresponding author on reasonable request. **B.** Detection of the *STAB2* splice-site variant in homozygous and heterozygous state in genomic DNA. **C.** Detection of an aberrant *STAB2* transcript, in comparison with wild-type, in cDNA derived from fibroblasts prepared from puromycin-treated cultures. Two sites in intron 54 gain recognition as donor and acceptor splice sites (blue arrows), and the intron 54 acceptor and intron 55 donor splice sites are lost (red arrows). **D.** Aberrations seen in the patient’s fibroblast cDNA were recapitulated in a minigene approach. Left panel: schematic of the Exontrap vector pET01 containing *STAB2* exon 55 and flanking intron sequences. Next panel: gel image of RT-PCR products in wild-type and mutation-containing minigenes, with an empty vector control and a no-template control. Next panels: schematic and Sanger traces of RT-PCR products demonstrating a major transcript that lacks exon 55 resulting in a frameshift and truncation (p.Gly1919Profs*164) and minor amounts of an intron 54-derived pseudo-exon resulting in a frameshift and truncation (p.Gly1919Glufs*24). All transcripts contain premature stop codons and are therefore predicted to undergo nonsense-mediated decay. **E, F.** Immunohistochemistry for stabilin-2 (clone 3.1) on a liver biopsy from a human control shows strong sinusoidal staining (E), whereas stabilin-2 signal is absent in the proband’s mother (F) demonstrating stabilin-2 deficiency in this individual who is homozygous for the NM_017564:c.5755-1G>C variant. Both sections were stained on the same slide in a single run. Scale bars, 100 µm.

Exome sequencing revealed several large chromosomal regions of homozygosity in the proband, in line with the self-reported close parental consanguinity and it did not identify pathogenic or likely pathogenic variants in established thrombophilia-associated genes. Exome sequencing identified a homozygous *STAB2* splice-site variant in the proband, c.5755-1G>C (Figure 1B), affecting the invariant –1 nucleotide of the canonical splice-acceptor site immediately upstream of exon 55. The variant caused exon 55 skipping in all detectable *STAB2* transcripts and inclusion of intron 54 sequence in minor amounts of transcript, as seen in fibroblast-derived RNA and in a minigene splicing system (1C,D), in agreement with *in silico* prediction (Supplemental Figure 1). The abnormal transcripts are predicted to contain premature termination codons and therefore to be subject to nonsense-mediated decay.

Segregation analysis showed that the proband’s healthy mother (I-2) and one healthy brother (II-2), both without a history of VTE, were also homozygous for this splice-site variant. Stabilin-2 deficiency could not be demonstrated on the protein level in cultured fibroblasts or peripheral blood because of low *STAB2* expression in these tissues. However, immunohistochemistry for stabilin-2 on a liver biopsy from the proband’s mother revealed no detectable signal, whereas control human liver routinely included as a positive control in the diagnostic staining protocol and stained on the same slide, showed strong labelling of liver sinusoidal endothelial cells (Fig. 1E,F). The homozygous *STAB2* splice-site variant thus results in loss of detectable stabilin-2 protein. Consequently, the observation of approximately 900-to 2000-fold elevated serum HA levels in three homozygous carriers strongly supports severe impairment of stabilin-2-dependent HA clearance. Elevated HA levels were repeatedly observed over a period of more than 10 years in the proband’s mother.

Plasma FVIII levels and VWF activity were elevated in all three homozygous individuals, and VWF antigen was elevated in two of the three individuals, supporting the concept that stabilin-2 contributes to the turnover of VWF and FVIII in the circulation. Despite similarly elevated HA, VWF, and FVIII levels, the two additional homozygous family members had no history of VTE, indicating variable clinical expression of the phenotype.

Fibrinogen levels were within the reference range in the examined homozygous individuals. Elevated FVIII levels were accompanied by abnormal thrombin generation potential in the proband’s healthy mother (Supplemental Table 1), homozygous for *STAB2* c.5755-1G>C, consistent with a procoagulant plasma phenotype. Detailed analyses of blood coagulation, blood groups, and blood counts are shown in Table 1. Heterozygous carriers displayed plasma VWF and FVIII and serum HA levels within the reference ranges.

**Table 1.** Patient characteristics.

| Individual<br>(Fig. 1A) | Reference<br>ranges | I-2 | II-1 | II-2 | II-3 | I-1 |
| --- | --- | --- | --- | --- | --- | --- |
| *STAB2*<br>splice-site<br>variant |  | Hom | Hom | Hom | Het | Het |
| VTE history |  | None | 8 episodes | None | None | None |
| Blood group |  | 0 | 0 | 0 | 0 | 0 |
| Factor<br>VIIIc, % | 70-200 | 223 | 255 | 321 | 142 | 111 |
| VWF-Ag, % | 70-150 | 187 | 141 | 193 | 132 | 75 |
| VWF<br>activity, % | 70-150 | 160 | 171 | 171 | 145 | 71 |
| Fibrinogen,<br>g/L | 1.5-4.5 | 2.9 | 1.8 | 2.3 | 2.2 | 3.1 |
| Quick, % | 70-130 | 74 | 79 | 103 | 101 | 75 |
| INR | 0.9-1.15 | 1.15 | 1.10 | 0.98 | 0.99 | 1.12 |
| aPTT, sec | 15-30 | 22 | 25.3 | 23 | 26.2 | 27.2 |
| D-Dimer,<br>µg/mL | 0-0.5 | 0.47 | 0.2 | 0.2 | 0.2 | 0.2 |
| HA, µg/L | 0-195 | 169281 | 393000 | 362013 | nd | 132 |
| HA <100<br>kDa, µg/L | <100 | 1783 | nd | 6130 | nd | nd |
| HA <10<br>kDa, µg/L | <100 | 646 | nd | 6058 | nd | nd |

Abbreviations: aPTT, activated partial thromboplastin time; HA, hyaluronic acid; Het, heterozygous; Hom, homozygous; INR, international normalized ratio; nd, not determined; Quick, prothrombin time reported as Quick value; VTE, venous thromboembolism; VWF, von Willebrand factor; WT, wild-type.
To protect participant privacy, results of more distant relatives are not shown individually: four heterozygous carriers and one non-carrier had serum HA levels within the reference range (33–76 µg/L) and Quick, INR, and
aPTT values within the reference ranges, where determined; individual data are available from the corresponding author on reasonable request.

Stabilin-2 deficiency was not associated with abnormal blood, serum, or plasma viscosity in the examined individual, nor in stabilin-2-knockout mice (Supplemental Table 2). The factor V Leiden mutation and the common prothrombin 20210G>A variant were not present in this family.

## Discussion

This study documents markedly elevated circulating HA levels in humans homozygous for a disruptive *STAB2* splice-site variant that results in stabilin-2 protein abrogation. These findings are consistent with an important role for stabilin-2 in systemic HA clearance in humans. The observation that one adult homozygous individual remained clinically well and had preserved fertility indicates that chronically elevated circulating HA can be tolerated without an overt syndromic phenotype in at least some affected adults. This further suggests that loss of stabilin-2-dependent HA clearance alone is compatible with normal human development and homeostasis. This hypothesis is supported by the very recent report of an adult male individual who carried a homozygous *STAB2* frameshift mutation predicting stabilin-2 deficiency together with a rare heterozygous *STAB1* mutation. This individual presented with mild chronic proteinuria and elevated blood pressure but without history of VTE. A kidney biopsy disclosed the rare type of chronic kidney disease, collagenofibrotic glomerulopathy, in this subject. Serum HA levels were highly elevated. In line with our observations here, it was hypothesized that stabilin-2 deficiency *per se* might go unnoticed clinically, but when combined with a pathogenic *STAB1* mutation, causes collagenofibrotic glomerulopathy.^33^ Biallelic *STAB2* LoF variants are rare in the general population. In gnomAD v4.1.0, only very few homozygous LoF carriers are observed, and comparable numbers of compound heterozygotes may be expected.^34^ Altogether, LoF variants appear mildly underrepresented, which may be compatible with some increase in morbidity but does not suggest a uniformly severe phenotype.

The glycosaminoglycan HA serves as a morphogenic matrix during development and tissue homeostasis, and cell-bound HA plays a role in the extravasation of lymphocytes and neutrophils from blood capillaries as well as macrophage entry into lymphatic capillaries. Because HA function depends on molecular size, the finding that all measured HA size fractions were markedly elevated is of interest, although the biological interpretation of this pattern remains limited and represents a constraint of the present study.

Stabilin-2-expressing cells bind and internalize human VWF and FVIII in a VWF-dependent manner.^7^ Stabilin-2 deficiency is therefore expected to result in elevated levels of VWF and FVIII. Plasma FVIII levels are a significant independent risk factor for VTE in population studies and are influenced by both VWF levels and ABO blood group.^35,36^ High FVIII levels have also been associated with an increased risk of childhood VTE.^37,38^ In the present family, elevated VWF and FVIII levels in all homozygous carriers provide the more plausible link to thrombosis susceptibility than HA itself.

The findings are consistent with the interpretation that impaired stabilin-2-dependent clearance contributed to elevated VWF and FVIII levels and thereby may have lowered the threshold for VTE in the proband, whose clinical course also included major acquired thrombotic risk factors related to severe intestinal disease, long-term parenteral nutrition, repeated central venous access, and septic episodes. This interpretation is compatible with the increased thrombotic burden and altered composition of venous thrombi reported in stabilin-2-deficient mice upon DVT induction.^39^ The present study does not support a direct prothrombotic role of elevated HA itself. Instead, the phenotype is more consistent with incomplete penetrance of a thrombosis susceptibility state.

Elevated HA serum levels have previously been associated with beneficial effects in some experimental settings, including reduced atherogenesis, reduced metastasis, and improved healthspan in mice.^10,40,41^ In human hyaluronidase deficiency, serum HA levels are elevated approximately 10-fold above normal without associated VTE.^42^ These observations further support caution in assuming that elevated circulating HA is intrinsically harmful or prothrombotic. One UK Biobank study identified associations for the clearance of 41 plasma proteins for *STAB2*,^14^ of which only three were addressed in our study.

Taken together, this family-based study supports the conclusion from population studies that stabilin-2 dysfunction may represent a thrombosis susceptibility factor relevant to VTE evaluation. Markedly elevated circulating HA may serve as a biochemical clue to impaired stabilin-2-dependent clearance in individuals with suspected disruptive *STAB2* variants, but the present data do not support HA as an isolated marker of VTE risk. The family phenotype instead suggests incomplete penetrance and variable expressivity, as commonly observed with inherited thrombophilia, with markedly abnormal biomarker profiles but no uniform clinical phenotype among homozygous carriers. The low number of affected individuals remains a limitation, but the present findings help define the clinical significance of stabilin-2 deficiency in humans.

## Data Availability

The data of this study are available from the corresponding author upon reasonable request.

## Supplemental Material

Supplemental Material.pdf

Table S1

Table S2

Figures S1

