## Supplemental Material for "Procoagulant plasma in three individuals with stabilin-2 deficiency"

**Short title:** Stabilin-2 deficiency

Rüdiger Adam[1], Alexander J. Jordan[2], Taras Valovka[3], Carla S. M. Seidl[3], Karin Kreidl[3], Carl-Erik Dempfle[4], Dorota Garczarczyk-Asim[3], Annelie Siegemund[5], Stefanie Nittka[6], Robert Müller[7,12], Petra Bürgisser[13], Ruben Bierings[13], Christoph Brochhausen-Delius[17], Krystin Krauel[14], Alice Assinger[15,16], Lea Berger[8], Eray Yagmur[9], Anja Schmaus[10], Werner Streif[3], Cyrill Géraud[8,*], Thomas Müller[3,*], Andreas R. Janecke[3,11,*]

[1] Department of General Pediatrics, DONAUISAR Klinikum Deggendorf and Faculty of Medicine, Regensburg University, 93053 Regensburg, Germany

[2] Department of General Pediatrics, University Medical Center and Medical Faculty Mannheim, Heidelberg University, 68167 Mannheim, Germany.

[3] Department of Pediatrics I, Medical University of Innsbruck, 6020 Innsbruck, Austria.

[4] IMD Coagulation Center, 61867 Mannheim, Germany.

[5] MVZ Limbach Magdeburg, Center of Coagulation Disorders and Vascular Diseases, 39104 Magdeburg, Germany.

[6] Institute for Clinical Chemistry, University Medical Center and Medical Faculty Mannheim, University of Heidelberg, 68167 Mannheim, Germany.

[7] MVZ Labor Ravensburg, 88212 Ravensburg, Germany.

[8] European Center for Angioscience and Section of Clinical and Molecular Dermatology, Department of Dermatology, Venereology, and Allergology, University Medical Center and

Medical Faculty Mannheim, Heidelberg University, 68167 Mannheim, Germany.

[9] Institute of Laboratory Medicine, Westpfalz Hospital, 67655 Kaiserslautern, Germany.

[10] European Center for Angioscience (ECAS), Medical Faculty Mannheim, Heidelberg University, 68167 Mannheim, Germany.

[11] Institute of Human Genetics, Medical University of Innsbruck, 6020 Innsbruck, Austria.

[12] Private University of the Principality of Liechtenstein, 9495 Triesen, Liechtenstein.

[13] Hematology, Erasmus University Medical Center, 3015 AA Rotterdam, Netherlands.

[14] Department of Cardiology, Haemostaseology, and Medical Intensive Care, University Medical Center and Medical Faculty Mannheim, Heidelberg University, 68167 Mannheim, Germany.

[15] Institute of Vascular Biology and Thrombosis Research, Department of Physiology and Pharmacology, Medical University of Vienna, 1090 Vienna, Austria.

[16] Ludwig Boltzmann Institute for Cardiovascular Research, 1090 Vienna, Austria.

[17] Institute of Pathology, University Medical Center and Medical Faculty Mannheim, Heidelberg University, 68167 Mannheim, Germany.

*Equal contribution

**Correspondence:**

Andreas R. Janecke, M.D. & Thomas Müller, M.D., Medical University of Innsbruck, Department of Pediatrics I, Anichstrasse 35, A-6020 Innsbruck, Austria; Phone: +43 512 504 23600; &, and Cyrill Geraud, European Center for Angioscience and Section of Clinical and Molecular Dermatology, Department of Dermatology, Venereology, and Allergology, University Medical Center and Medical Faculty Mannheim, Heidelberg University, 68167 Mannheim, Germany,

**Supplemental Table 1. Thrombin generation on ST Genesia in individual I-2**

| Thrombin generation (TG) (-TM), unit | Reference (Median [25p-75p] 95% CI)* | Individual I-2 (Fig. 1A) |
|---|---|---|
| Lag time, min | 2.4 [2.3–2.6] 2.1–3.1 | 3.33 |
| Peak height, nmol/L | 172 [134–227] 107–268 | 241.1 |
| Time-to-peak, min | 5.1 [4.8–5.4] 4.5–6.0 | 6.18 |
| ETP, nmol/L.min | 1167 [1010–1274] 867–1570 | 1389 |
| Velocity Index, nM/min | 85 [67–130] 50–152 | 106.3 |
| **(+TM), unit** | | |
| Lag time, min | 2.5 [2.4–2.7] 2.2–3.3 | 3.32 |
| Peak height, nmol/L | 153 [118–203] 79–224 | 207.4 |
| Time-to-peak, min | 4.7 [4.5–4.9] 4.3–5.6 | 5.58 |
| ETP, nmol/L.min | 792 [610–969] 406–1136 | 1104 |
| Velocity Index, nM/min | 95 [65–131] 48–149 | 120.0 |
| ETP inhibition ratio | 34 [25–43] 20–53 | 20.57 |

TM, Thrombomodulin; CI, confidence interval; ETP, endogenous thrombin generation

* Reference values are taken from a total of 26 healthy female subjects with a median age of 17 years, not using oral contraceptives, as reported[1].

**Supplemental Table 2. Blood, plasma and serum viscosity in individuals with stabilin-2 deficiency and in the stabilin-2 knockout mouse**

| Individual | Material | Result (mPa•s) | Reference |
|---|---|---|---|
| I-2 | blood | 4.22 | 3.9 - 4.7 |
| II-2 | Plasma | 1.39 | 1.2 - 1.9 |
| II-1 | Plasma | 1.67 | 1.2 - 1.9 |
| I-2 | Plasma | 1.51 | 1.2 - 1.9 |
| I-2 | Serum | 1.36 | 1.1 - 1.7 |

| | | |
|---|---|---|
| Stab2KO mouse male | Plasma | 1.03 |
| Stab2KO mouse female | Plasma | 1.07 |
| Wildtype mouse male | Plasma | 0.96 |
| Wildtype mouse female | Plasma | 0.99 |

**Supplemental Figure S1. In silico analysis of the *STAB2* splice-site mutation**

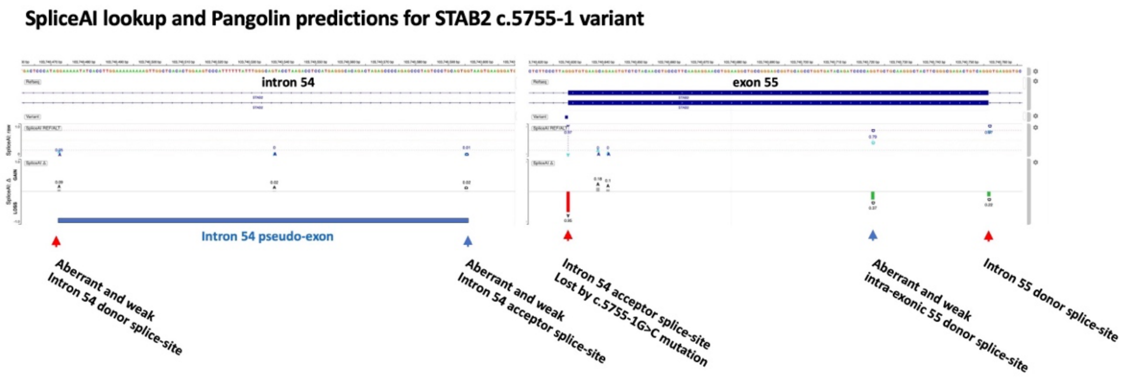

Two sites in intron 54 gain recognition as donor and acceptor splice sites (blue arrows), and the intron 54 acceptor and intron 55 donor splice sites are lost (red arrows) are predicted consequences by the spliceAI lookup and the Pangolin tools as consequences resulting from the *STAB2* variant c.5755-1G>C, affecting a canonical splice-site https://spliceailookup.broadinstitute.org/#variant=chr12-103740629-G-C&hg=38&bc=basic&distance=500&mask=0&ra=0.
